# Prenatal cannabis exposure and childhood outcomes: Results from the ABCD study

**DOI:** 10.1101/2019.12.18.19015164

**Authors:** Sarah E. Paul, Alexander Hatoum, Jeremy D. Fine, Emma C. Johnson, Isabella Hansen, Nicole R. Karcher, Allison L. Moreau, Erin Bondy, Yueyue Qu, Ebony B. Carter, Cynthia E Rogers, Arpana Agrawal, Deanna M. Barch, Ryan Bogdan

**Author notes:** Author’s note: Correspondence concerning this article should be addressed to Ryan Bogdan, address: CB 1125 Psychological and Brain Sciences Bldg Room 453, Washington University in St. Louis, One Brookings Drive, St. Louis, MO 63130, USA) and Sarah Paul, address: CB 1125 Psychological and Brain Sciences Bldg Room 453, Washington University in St. Louis, One Brookings Drive, St. Louis, MO 63130, USA.

## Abstract

**Importance:** In light of increasing cannabis use among pregnant women, the Surgeon General of the United States issued an advisory against the use of marijuana during pregnancy on August 29^th^, 2019.

**Objective:** To determine whether cannabis use during pregnancy is associated with adverse outcomes among offspring.

**Design:** Cross-sectional analysis of the baseline session of the ongoing longitudinal Adolescent Brain and Cognitive Development (ABCD) study.

**Setting:** Data were collected from 22 sites across the United States between 2016 and 2018.

**Participants:** Children ages 9-11 (n=11,489) and their parent or caregiver.

**Exposure:** Prenatal marijuana exposure prior to and following maternal knowledge of pregnancy.

**Main Outcomes and Measures:** Child psychopathology symptomatology (i.e., psychotic-like experiences and internalizing, externalizing, attention, thought, and social problems), cognition, sleep, birth weight, gestational age at birth, body mass index (BMI), and brain structure (i.e., total intracranial volume, white matter volume, gray matter volume). Covariates included familial (e.g., income, familial psychopathology), pregnancy (e.g., prenatal vitamin use, whether the pregnancy was planned), and child (e.g., birth weight, substance use) variables.

**Results:** Among 11,489 children (age 9.9±0.6 years; 47.78% female), 655 (5.70%) were prenatally exposed to cannabis in total. Marijuana use prior to (n=648; 5.64%) and following (n=242; 2.11%) maternal knowledge of pregnancy were associated with increased offspring psychopathology characteristics (i.e., psychotic-like experiences and internalizing, externalizing, attention, thought, social, and sleep problems) and BMI as well as reduced cognition, birth weight, and brain structure (i.e., total white and gray mater volumes; all *ps*_*fdr*_<.007), but not gestational age at birth. Exposure following maternal knowledge of pregnancy remained significantly associated with psychopathology, cognition, and birth weight outcomes when including potentially confounding variables (all *p*s<0.046). All associations with exposure prior to maternal knowledge of pregnancy were nonsignificant when considering potentially confounding variables (all *p*s>0.06).

**Conclusions and Relevance:** Prenatal cannabis exposure, and its correlated factors, may increase risk for psychopathology and reduced cognition during middle childhood as well as reduced birthweight. Consistent with recent recommendations by the Surgeon General, marijuana use during pregnancy should be discouraged.

## INTRODUCTION

Alongside increasingly permissive sociocultural attitudes and laws surrounding cannabis,^1^ past month use among pregnant women increased by 106% from 2002(3.4%) to 2017(7.0%) in the United States.^2^ Tetrahydrocannabinol (THC), the psychoactive component of cannabis, crosses the placenta^3^ and interfaces with the endocannabinoid system, which critically influences neural development.^3-5^ Thus, it is plausible that cannabis use during pregnancy may impact offspring. Indeed, the alarming increase of cannabis use among pregnant mothers,^2,6-8^ and evidence linking prenatal exposure to adverse outcomes in children^9-13^ prompted the Surgeon General of the United States to release an advisory against cannabis use during pregnancy and breastfeeding on August 29^th^ 2019.^14^

Despite the increasing prevalence of prenatal cannabis exposure, there have been relatively few investigations of its association with child outcomes. These studies have found evidence linking prenatal marijuana exposure to reduced birth weight^15^ and cognition^16,17^ as well as heightened risk for premature birth,^18^ psychopathology (i.e., psychosis, internalizing, externalizing)^19-21^ and sleep problems^22^ among children. However, there has been limited cross-study replication,^e.g.,23-25^ and studies have typically been unable to account for other potential confounding factors (e.g., maternal education, prenatal vitamin usage, child substance use, familial risk), leaving it possible that associations between prenatal marijuana exposure and negative outcomes among children may not be independent of them. Indeed, the National Academies of Sciences, Engineering, and Medicine recently concluded that only reduced birth weight has been robustly linked to prenatal cannabis exposure.^13^

Using data from the Adolescent Brain Cognitive Development (ABCD) study (data release 2.0.1) of 11,875 children, we test whether prenatal marijuana exposure before and after maternal knowledge of pregnancy is associated with psychopathology (i.e., internalizing, externalizing, attention, thought and social problems, psychotic-like experiences), sleep, cognition, birth weight, premature birth, body mass index (BMI), and gross brain structure (i.e., white and gray matter volumes as well as intracranial volume). This represents a comprehensive extension of our prior investigation of prenatal cannabis exposure and psychotic-like experiences among the initial ABCD data release (n=4,361).^26^ We hypothesized that prenatal marijuana exposure before and after maternal knowledge of pregnancy would be associated with greater risk for psychopathology, premature birth, and sleep problems, as well as reduced cognition, birth weight, BMI, and brain structure metrics when not considering potentially confounding variables. However, given that endocannabinoid receptors are not expressed in the fetus until 5-6 weeks^9,27,28^ which roughly corresponds to when, in this study, mothers learned that they were pregnant (6.91 ± 6.75 weeks), we hypothesized that these associations would only be robust to the inclusion of potentially confounding covariates when marijuana use occurred following maternal knowledge of pregnancy.

## METHODS

### Participants

Data came from children (n=11,875; *M*_*age*_ *=* 9.9±0.6 years; 47.85% girls; 74.13% White), born between 2005 and 2009 to 9,987 mothers through 10,801 pregnancies, who completed the baseline session of the ongoing longitudinal Adolescent Brain Cognitive Development (ABCD) study (data release 2.0.1; https://abcdstudy.org/).^29^ The study includes a family-based design in which twin (n=2,108), triplet (n=30), non-twin siblings (n=1,589), and singletons (n=8,148) were recruited. All parents/caregivers (85.3% biological mothers)^a^ and children provided written informed consent and verbal assent, respectively, to a research protocol approved by the institutional review board at each data collection site (n=22)^b^ throughout the United States (https://abcdstudy.org/sites/abcd-sites.html). For our analyses, only data from participants with non-missing data on prenatal cannabis exposure were included (n=11,489 [87.6% of parent/caregivers were biological mothers]; *M*_*age*_ *=* 9.9±0.6 years; 47.78% girls; 74.76% self-reported White; 2,072 twins, 30 triplets, 1,523 non-twin siblings, 7,864 singletons; **Table 1**).

**Table 1.** ABCD Sample Characteristics.

| Variable (N) | Prenatal Cannabis Exposure Post-Knowledge of Pregnancy (N=242) | No Prenatal Cannabis Exposure Post-Knowledge of Pregnancy |  |  | Total (11489) |
| --- | --- | --- | --- | --- | --- |
|  |  | Prenatal Cannabis Use Before Knowing (N=413) | No Prenatal Use (N=10834) | Total (N=11247) |  |
| Child Variables |  |  |  |  |  |
| Age in years (11,489) | 9.83 ± 0.62 | 9.89 ± 0.63 | 9.92 ± 0.62 | 9.91 ± 0.62 | 9.91 ± 0.62 |
| Gender, female (11,486) | 131 (54.13%) | 194 (46.97%) | 5164 (47.66%) | 5358 (47.64%) | 5489 (47.78%) |
| Birthweight in oz. (11,113) | 108.80 ± 23.84 | 112.6 ± 22.57 | 112.30 ± 23.45 | 112.30 ± 23.42 | 112.20 ± 23.43 |
| Race/Ethnicity (11,489)<sup>a</sup> |  |  |  |  |  |
| White | 144 (59.50%) | 242 (58.60%) | 8203 (75.72%) | 8445 (75.09%) | 8589 (74.76%) |
| Black | 110 (45.45%) | 172 (41.65%) | 2094 (19.33%) | 2266 (20.15%) | 2376 (20.68%) |
| Asian | 5 (2.07%) | 13 (3.15%) | 692 (6.39%) | 705 (6.27%) | 710 (6.18%) |
| Pacific Islander | 1 (0.41%) | 5 (1.21%) | 63 (0.58%) | 68 (0.60%) | 69 (0.60%) |
| Native American | 14 (5.79%) | 17 (4.12%) | 3.13 (3.17%) | 360 (3.20%) | 374 (3.26%) |
| Hispanic | 34 (14.05%) | 89 (21.55%) | 2215 (20.44%) | 2304 (20.49%) | 2338 (20.35%) |
| Other | 8 (3.31%) | 34 (8.23%) | 722 (6.66%) | 756 (6.72%) | 764 (6.65%) |
| Child Lifetime Substance Exposure |  |  |  |  |  |
| Alcohol Try (11,482) | 52 (21.49%) | 113 (27.36%) | 2417 (22.31%) | 2530 (22.49%) | 2582 (22.49%) |
| Tobacco Try (11,482) | 10 (4.13%) | 12 (2.91%) | 98 (0.90%) | 110 (0.98%) | 120 (1.05%) |
| Marijuana Try (11,482) | 3 (1.24%) | 2 (0.48%) | 9 (0.08%) | 11 (0.10%) | 14 (0.12%) |
| Pregnancy and Family Variables |  |  |  |  |  |
| Unplanned Pregnancy (11,377) | 170 (70.25%) | 111 (26.88%) | 3865 (35.67%) | 4166 (37.04%) | 4336 (37.74%) |
| Maternal Age at Birth (11,336) | 25.33 ± 6.36 | 25.39 ± 5.90 | 29.71 ± 6.14 | 29.55 ± 6.18 | 29.47 ± 6.22 |
| Prenatal Vitamin Use (11,236) | 178 (73.55%) | 392 (94.92%) | 10172 (93.89%) | 10564 (93.93%) | 10742 (93.50%) |
| Week Learned Pregnancy (10,375) | 8.30 ± 7.39 | 7.84 ± 6.48 | 6.85 ± 6.74 | 6.88 ± 6.74 | 6.91 ± 6.75 |
| Maternal Education: Years (10974) | 13.68 ± 2.01 | 14.27 ± 2.23 | 15.27 ± 2.62 | 15.24 ± 2.61 | 15.21 ± 2.61 |
| Household Inc (10,507) |  |  |  |  |  |
| \$0-\$49,999k | 136 (56.20%) | 204 (49.39%) | 2764 (25.51%) | 2968 (26.39%) | 3104 (27.02%) |
| \$50k-\$74,999k | 32 (13.22%) | 54 (13.08%) | 1368 (12.63%) | 1422 (12.64%) | 1454 (12.67%) |
| \$75k-\$99,999k | 29 (11.98%) | 37 (8.96%) | 1445 (13.34%) | 1482 (13.18%) | 1511 (13.15%) |
| \$100k-\$199,999k | 21 (8.68%) | 60 (14.53%) | 3136 (28.95%) | 3196 (28.42%) | 3217 (28.00%) |
| \$200k or more | 2 (0.83%) | 21 (5.08%) | 1198 (11.06%) | 1219 (10.84%) | 1221 (10.63%) |
| Family History of Psychopathology<sup>b</sup> |  |  |  |  |  |
| Psychosis (11,205) | 15 (6.20%) | 23 (5.57%) | 223 (2.06%) | 246 (2.19%) | 261 (2.27%) |
| Depression (11,329) | 139 (57.44%) | 194 (46.97%) | 3280 (30.28%) | 3474 (30.89%) | 3613 (31.45%) |
| Anxiety (11,150) | 58 (23.97%) | 101 (24.46%) | 1216 (11.22%) | 1317 (11.71%) | 1375 (11.97%) |
| Antisocial Behavior (11,320) | 123 (50.83%) | 151 (36.56%) | 1146 (10.58%) | 1298 (11.54%) | 1421 (12.37%) |
| Mania (11,138) | 47 (19.42%) | 46 (11.14%) | 493 (4.55%) | 539 (4.79%) | 586 (5.10%) |
| Prenatal Substance Exposure Before Knowing of Pregnancy |  |  |  |  |  |
| Alcohol (11,072) | 135 (55.79%) | 255 (61.74%) | 2391 (22.07%) | 2646 (23.53%) | 2781 (24.21%) |
| Tobacco (11,430) | 59 (65.70%) | 230 (55.69%) | 1117 (10.31%) | 1347 (11.98%) | 1506 (13.11%) |
| Cannabis (11,489) | 235 (97.11%) | 413 (100.00%) | 0 (0.00%) | 413 (3.67%) | 648 (5.64%) |
| Prenatal Substance Exposure After Knowing of Pregnancy |  |  |  |  |  |
| Alcohol (11,446) | 63 (26.03%) | 13 (3.15%) | 214 (1.98%) | 227 (2.02%) | 290 (2.52%) |
| Tobacco (11,458) | 114 (47.11%) | 60 (14.53%) | 392 (3.62%) | 452 (4.02%) | 566 (4.93%) |
| Cannabis (11,489) | 242 (100.00%) | 0 (0.00%) | 0 (0.00%) | 0 (0.00%) | 242 (2.11%) |
| Child Primary Outcomes of Interest |  |  |  |  |  |
| Psychotic-like Experiences (11,477) | 3.58 ± 3.98 | 3.17 ± 3.67 | 2.57 ± 3.52 | 2.59 ± 3.53 | 2.61 ± 3.54 |
| Internalizing Sx (CBCL) (11,483) | 7.83 ± 7.39 | 6.41 ± 6.08 | 4.88 ± 5.38 | 4.94 ± 5.41 | 5.00 ± 5.48 |
| Externalizing Sx (CBCL) (11,483) | 10.06 ± 9.15 | 6.44 ± 7.36 | 4.13 ± 5.47 | 4.22 ± 5.56 | 4.34 ± 5.72 |
| Attention Sx (CBCL) (11,483) | 5.53 ± 4.32 | 4.23 ± 4.04 | 2.80 ± 3.36 | 2.86 ± 3.40 | 2.91 ± 3.44 |
| Thought Sx (CBCL) (11,483) | 3.28 ± 3.42 | 2.24 ± 2.79 | 1.53 ± 2.08 | 1.56 ± 2.12 | 1.59 ± 2.17 |
| Social Sx (CBCL) (11,483) | 3.36 ± 3.19 | 2.31 ± 2.68 | 1.53 ± 2.18 | 1.56 ± 2.21 | 1.59 ± 2.25 |
| Cognition Composite (11,249) | 81.65 ± 8.62 | 84.62 ± 9.41 | 86.49 ± 9.09 | 86.42 ± 9.11 | 86.32 ± 9.12 |
| Sleep problems (SDSC) (11,363) | 41.94 ± 13.52 | 38.88 ± 9.79 | 36.14 ± 8.13 | 36.24 ± 8.21 | 36.36 ± 8.39 |
| BMI (11,462) | 20.72 ± 5.89 | 19.73 ± 4.31 | 18.76 ± 4.18 | 18.79 ± 4.19 | 18.84 ± 4.24 |
| Gestational age at birth: weeks (11,414) | 39.31 ± 1.81 | 39.19 ± 2.06 | 39.07 ± 2.20 | 39.07 ± 2.20 | 39.08 ± 2.92 |
| Total Gray Matter Volume (11,407) | 9.98 ± 0.90 | 10.22 ± 0.95 | 10.39 ± 1.00 | 10.38 ± 1.00 | 10.38 ± 1.00 |
| Total White Matter Volume (10,790) | 8.71 ± 0.95 | 8.92 ± 1.07 | 9.04 ± 0.99 | 9.04 ± 1.00 | 9.03 ± 1.00 |
| Total Intracranial Volume(11,410) | 9.89 ± 0.87 | 10.06 ± 0.94 | 10.16 ± 1.00 | 10.16 ± 1.00 | 10.15 ± 1.00 |

### Measures

All measures are described in the **Supplement** and **Table 1**.

### Prenatal Cannabis Exposure

Child prenatal marijuana exposure before (n=648) or after (n=242) maternal knowledge of pregnancy were coded as separate dichotomous variables based upon parent/caregiver^a^ retrospective report.

### Psychotic-like experiences

The total score of the *Prodromal Questionnaire-Brief Child Version* (PQ-BC)^30,31^ was used to assess child-reported psychotic-like experiences (PLEs). Higher scores indicate more PLEs.

### Internalizing, Externalizing, Attention, Thought, and Social Problems

The *Child Behavior Checklist* (CBCL)^32^ was used to assess broad spectrum internalizing (INT) and externalizing (EXT) as well as attention (ATN), thought (THT), and social (SOC) problems in children according to parent/caregiver report. Higher scores are reflective of more problems.

### Sleep Problems

The total score of the *Sleep Disturbance Scale for Children* (SDSC)^33^ was used to assess parent/caregiver-reported sleep problems in children. Higher scores reflect more sleep problems.

### Cognition

The *National Institutes of Health Toolbox Cognition Battery–Total Cognition Composite*^*34*^ indexed child cognitive ability. Higher scores are reflective of greater cognitive performance.

### Gestational Age at Birth, Birth Weight and Body Mass Index

A parent/caregiver retrospectively reported their child’s gestational age at birth in weeks and birth weight. BMI was calculated using measured height and weight at the baseline assessment.

### Brain Structure: Total Intracranial, White Matter, and Gray Matter Volumes

Freesurfer v5.3 was used to estimate total intracranial volume (ICV), gray matter volume (GMV), and white matter volume (WMV).

### Covariates

The following variables were dummy coded: *race/ethnicity, family history of psychopathology, prenatal exposure to tobacco or alcohol before or after maternal knowledge of pregnancy, unplanned pregnancy, prenatal vitamin usage, child alcohol try, child tobacco try, child self-reported gender*, and *twin/triplet status. Annual household income* was treated as a 5-level categorical variable. The following continuous covariates were included: *birthweight, maternal age at birth, gestational age when pregnancy was discovered (weeks), child age*, and *maternal education* (years). Total intracranial volume was further included as a covariate only in models with gray and white matter volumes as outcomes. Polygenic scores (PGS) for schizophrenia, educational attainment, birthweight, and cannabis use as well as ancestrally-informative principal components (n=10) were included as covariates in *post-hoc* analyses within the genomically-confirmed European ancestry subsample (**Statistical Analyses; Supplement**).

Non-prevalent substance use among children (n=80) or by women while they were pregnant (n=185) as well as extreme premature birth (n=148), and non-biological mother parent/caregiver report (n=1,427) were not included as covariates; we conducted *post hoc* analyses excluding individuals on these variables (**Supplement**).

### Statistical Analyses

Continuous predictor and outcome variables were winsorized to minimize the influence of extreme values (i.e., any values ±3 SDs were set at the ±3 SD point). We used linear mixed-effects models with random intercept parameters to account for non-independence of site and family membership for all non-imaging analyses and scanner and family for all imaging analyses, with the lme4 package in Rv3.6.0.^35^

We examined the association between prenatal cannabis exposure before and after maternal knowledge of pregnancy (each coded 0/1 separately) and outcomes using three analytic approaches. *First*, we tested whether prenatal marijuana exposure prior to and after maternal knowledge were associated with outcomes of interest in independent nested mixed models with no fixed effect covariates. We adjusted for multiple testing using Benjamini Hochberg false discovery rate correction (fdr_BH_). *Second*, for any associations that were significant after multiple testing correction, we examined whether they were robust to the inclusion of potentially confounding covariates; here, we simultaneously entered prenatal marijuana use before and after maternal knowledge of pregnancy alongside the covariates described above (except genomic data). To address concerns of overfitting, we reran all analyses including only covariates that were significantly associated with the outcome (**Supplement**). We conducted *post hoc* analyses to examine subcomponents of significant associations (i.e., CBCL INT and EXT subscales, cognition subcomponents; **Supplement**). *Third* and finally, to account for the possible confounding effects of genomic liability to offspring outcomes not accounted for by familial history, we examined whether associations remained significant when accounting for polygenic scores (PGS) derived using summary statistics from genome-wide association studies (GWASs) of related outcomes (**Supplement)**.^36^ Here, we evaluated whether any associations robust to covariate inclusion were present in the subsample of individuals with genomically-confirmed European ancestry (n=4,644) to ensure that any differences could not be attributable to power reductions, before examining whether these associations were robust to the inclusion of GWAS-weighted PGS (i.e., schizophrenia,^37^ educational attainment,^38^ cannabis use,^39^ polygenic scores for PLEs; cannabis use for social problems; birth weight^40^ and cannabis use for birth weight; **Supplement)**.

## RESULTS

Among 11,489 children (mean [SD]: age, 9.9 [0.6] years; 5,488 [47.78%] girls), 655 (5.85%) were prenatally exposed to marijuana (**Table 1**). Of these, 413 were exposed only before maternal knowledge of pregnancy while 235 were exposed both before and after maternal knowledge, and 7 only after maternal knowledge. Mothers learned of their pregnancy at 6.91 ± 6.75 weeks. Rates of tobacco and alcohol use during pregnancy, prior to and following maternal knowledge of pregnancy, were higher than cannabis use (**Table 1**) and modestly correlated with prenatal marijuana exposure (r_s_s 0.11-0.34; **Supplemental eTable 1**).

### Prenatal Cannabis Exposure After Maternal Knowledge of Pregnancy

Prenatal marijuana exposure *<u>after</u>* maternal knowledge of pregnancy was significantly associated with higher PLEs, BMI and internalizing, externalizing, attention, thought, social, and sleep problems, as well as lower cognition, birth weight, and intracranial and gray and white matter volumes (all |*B*s|>0.27, all *ps*_*fdr*_<0.002; **Table 2**), but not gestational age at birth (*B*=0.14, *p*_*fdr*_*=*0.271). PLEs, birth weight, and cognition, as well as internalizing, externalizing, attention, thought, and social problems, remained significantly associated even after accounting for potentially confounding covariates (all |*B*s|>0.52, all *p*s<0.046), but sleep problems, BMI, and total intracranial, gray matter, and white matter volumes were not (all |*B*s|<0.87, *p*s>0.27; **Figure 1**; **Table 3**; **Supplemental eTables 2-8**). *Post hoc* analyses showed globally similar associations across subfacets of EXT, but not INT or cognition (**Supplemental eTable 9**).

**Table 2.** Mixed Effects Regression Results for Prenatal Cannabis Exposure Among 10,404-11,489 Children Without Inclusion of Potentially Confounding Covariates.

| Outcome | MJ exposure post-knowledge |  |  | MJ exposure pre-knowledge |  |  | N |
| --- | --- | --- | --- | --- | --- | --- | --- |
|  | B [95% CI] | p | pFDR | B [95% CI] | p | pFDR |  |
| Psychotic-like Experiences | 1.12 [.70, 1.5] | **2.21e-07** | **3.44e-07** | 0.795 [.53, 1.1] | **3.65e-09** | **7.30e-09** | 11477 |
| Internalizing (CBCL) | 2.42 [1.8, 3.1] | **7.00e-13** | **1.63e-12** | 1.78 [1.4, 2.2] | **2.27e-17** | **5.30e-17** | 11483 |
| Externalizing (CBCL) | 4.68 [4.0, 5.4] | **1.17e-42** | **1.64e-41** | 2.93 [2.5, 3.3] | **3.93e-43** | **5.50e-42** | 11483 |
| Attention (CBCL) | 2.37 [1.9, 2.8] | **8.89e-27** | **4.15e-26** | 1.71 [1.4, 2.0] | **2.59e-35** | **1.81e-34** | 11483 |
| Thought (CBCL) | 1.36 [1.1, 1.6] | **1.63e-25** | **5.71e-25** | 0.870 [.71, 1.0] | **7.75e-27** | **2.71e-26** | 11483 |
| Social (CBCL) | 1.54 [1.3, 1.8] | **3.59e-29** | **2.51e-28** | 0.984 [.82, 1.2] | **8.87e-31** | **4.14e-30** | 11483 |
| Body Mass Index | 1.31 [.80, 1.8] | **6.58e-07** | **9.21e-07** | 0.945 [.62, 1.3] | **8.76e-09** | **1.53e-08** | 11462 |
| Cognition Composite | -3.83 [-5.0, -2.7] | **1.87e-10** | **3.74e-10** | -2.08 [-2.8, -1.4] | **2.22e-08** | **3.45e-08** | 11249 |
| Sleep Problems | 4.04 [3.0, 5.0] | **1.89e-15** | **5.29e-15** | 2.88 [2.3, 3.5] | **9.29e-20** | **2.60e-19** | 11489 |
| Gestational Age at Birth | 0.142 [-.11, .40] | 0.271 | 0.271 | 0.076 [-.08, .23] | 0.334 | 0.334 | 11414 |
| Birth Weight | -3.39 [-5.4, -1.3] | **0.00123** | **0.00132** | -1.75 [-3.0, -.49] | **0.00648** | **0.00698** | 11113 |
| Intracranial Volume | -0.273 [-.39, -.15] | **7.37e-06** | **9.38e-06** | 0.157 [-.23, -.08] | **3.72e-05** | **4.34e-05** | 11024 |
| White Matter Volume | -0.298 [-.43, -.16] | **1.49e-05** | **1.74e-05** | -0.184 [-.27, -.10] | **1.92e-05** | **2.44e-05** | 10404 |
| Gray Matter Volume | -0.382 [-.51, -.25] | **7.77e-09** | **1.36e-08** | -0.230 [-.31, -.15] | **2.65e-08** | **3.71e-08** | 11020 |

**Note.** Linear mixed effect models were used to analyze the associations between pre- and post-knowledge of pregnancy use of cannabis (analyzed separately) and each outcome, nesting data by research site and family ID (non-imaging analyses) and scanner and family ID (imaging analyses). Psychotic-like experiences were assessed with the Prodromal Questionnaire Brief-Report Child Version; CBCL = Child Behavior Checklist; uncorrected Cognition Composite from the NIH Toolbox; Sleep problems from the Parent Sleep Disturbance Scale for Children. N's differ due to listwise deletion of missing data. All Bs are unstandardized. Values in [] represent 95% Confidence Intervals. *pFDR* = p-values after false discovery rate correction for multiple testing, calculated separately for pre- and post-knowledge cannabis exposure. Bolded values indicate significant associations.

**Table 3.** Associations Between Prenatal Cannabis Exposure When Including Potentially Confounding Covariates.

| Outcome | MJ exposure post-knowledge |  | MJ exposure pre-knowledge |  | N |
| --- | --- | --- | --- | --- | --- |
|  | B [95% CI] | p | B [95% CI] | p |  |
| Psychotic-like Experiences | 0.941 [.25, 1.6] | **0.00810** | -0.091 [-.48, .30] | 0.647 | 8165 |
| Internalizing (CBCL) | 1.15 [.08, 2.2] | **0.0359** | -0.053 [-.66, .55] | 0.862 | 8168 |
| Externalizing (CBCL) | 1.90 [.84, 3.0] | **4.63e-04** | 0.146 [-.45, .74] | 0.630 | 8168 |
| Attention (CBCL) | 1.14 [.44, 1.8] | **0.00136** | 0.205 [-.19, .60] | 0.305 | 8168 |
| Thought (CBCL) | 0.523 [.11, .94] | **0.0134** | 0.129 [-.10, .36] | 0.277 | 8168 |
| Social (CBCL) | 0.942 [.51, 1.4] | **2.00e-05** | 0.070 [-.17, .31] | 0.572 | 8168 |
| Body Mass Index | 0.456 [-.36, 1.3] | 0.275 | -0.022 [-.48, .44] | 0.925 | 8151 |
| Cognition Composite | -1.69 [-3.3, -.04] | **0.0457** | 0.887 [-.04, 1.8] | 0.0609 | 8000 |
| Total sleep problems | 0.867 [-.76, 2.5] | 0.296 | 0.370 [-.54, 1.3] | 0.428 | 8169 |
| Birth weight (oz) | -3.42 [-6.5, -.38] | **0.0278** | 0.871 [-.84, 2.6] | 0.318 | 8169 |
| Intracranial Volume | 0.010 [-.16, .18] | 0.912 | 0.025 [-.07, .12] | 0.614 | 7846 |
| White matter | -0.030 [-.15, .09] | 0.624 | -0.037 [-.10, .03] | 0.288 | 7431 |
| Gray matter | -0.020 [-.12, .08] | 0.711 | 0.037 [-.02, .10] | 0.212 | 7842 |

**Figure 1.**
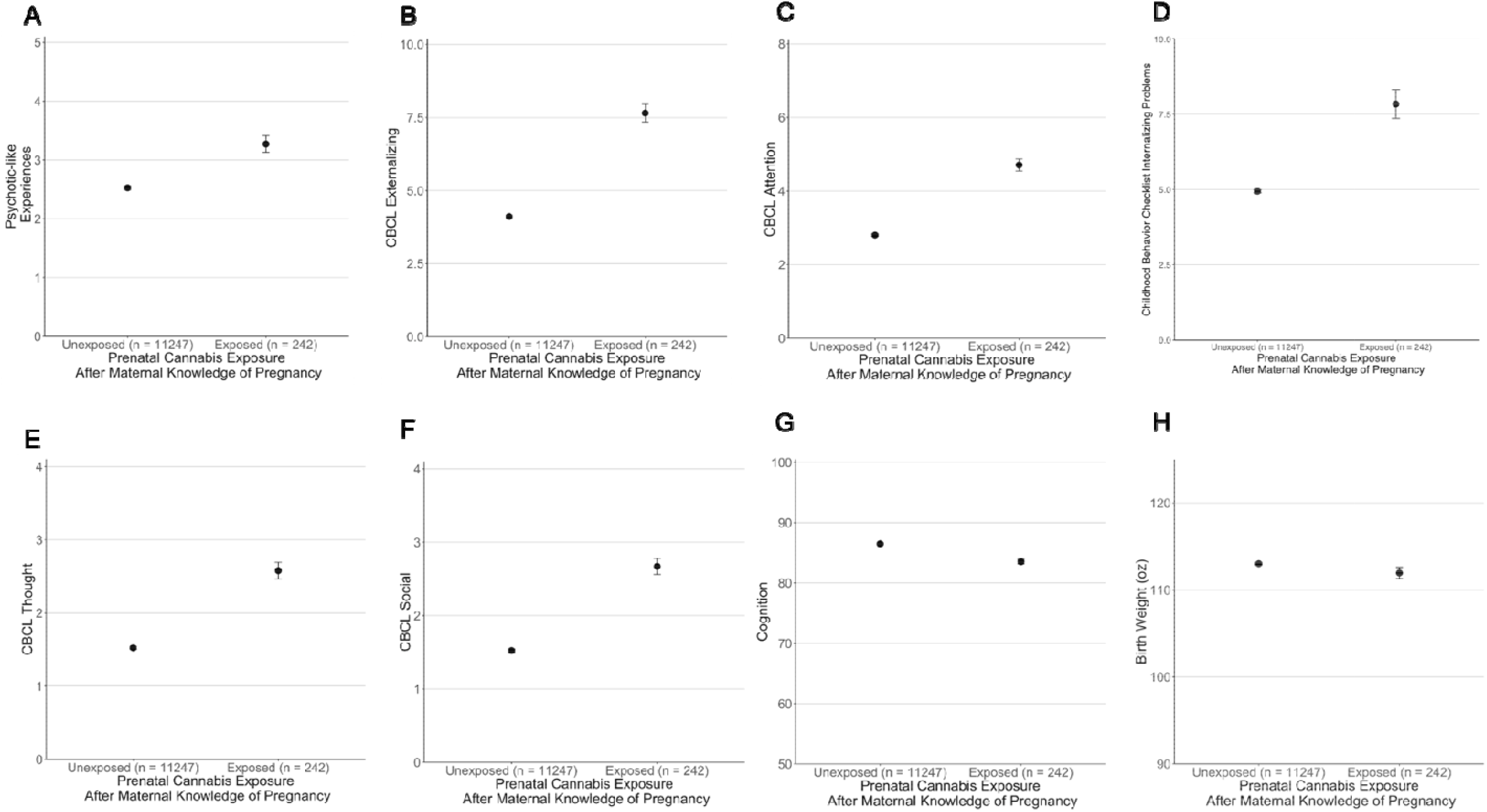
Prenatal Cannabis Exposure Post Maternal Knowledge of Pregnancy is Associated with Increased Psychopathology Risk (A-F), Reduced Cognition (G), and Reduced Birth Weight (H). Raw data values are plotted. As scales differ y axes are not directly comparable across panels. CBCL = Child Behavior Checklist. Statistics are presented in **Tables 2-3** and **Supplemental eTables 2-3**. Log--transforming data reduces differences in variability across groups and results in similar conclusions (**Supplemental eTable 5**).

Stepwise analyses of confounding covariates revealed that reductions in significance could not generally be attributed to a small collection of variables (**Supplement**). *Post Hoc Analyses* excluding: children who engaged in non-prevalent substance use, were exposed to other illicit substances prenatally, were born at extreme levels of prematurity, or whose biological mothers were not the parent/caregiver respondent revealed similar associations and consistent conclusions (**Supplemental eTable 10-13**). Finally, in the subsample of children with genomically-confirmed European ancestry (N=4,644; 3.94% reporting any prenatal exposure), PLEs, social problems, and birth weight remained significantly associated with prenatal exposure to cannabis after maternal knowledge of pregnancy even after accounting for covariates (all |*Bs*|>0.97, all *p*s<0.016; **Supplemental eTable 14**) and when further accounting for child PGS for schizophrenia, educational attainment, birth weight, and/or cannabis use (|*B*s|>0.95, all *p*s<0.018; **Supplemental eTables 15-18**).

### Prenatal Cannabis Exposure Prior to Maternal Knowledge of Pregnancy

Prenatal marijuana exposure *<u>prior</u> <u>to</u>* maternal knowledge of pregnancy was significantly associated with all outcomes (all |*B*s|>0.15, all *ps*_*fdr*_<7.0*10^−3^) except gestational age at birth (*B*=0.076, *p*_*fdr*_=0.334; **Table 2**). No associations remained significant after accounting for potentially confounding covariates, but a nominal trend for increased (directionally opposite to results without covariates) cognition emerged (**Table 3; Supplemental eTables 2-8**). Stepwise analysis of confounding covariates revealed that no small group of covariates was responsible for attenuating these associations (but see Attention and Thought problems; **Supplemental eResults**).

## DISCUSSION

Prenatal cannabis exposure following maternal knowledge of pregnancy is associated with elevated psychopathology risk and reduced cognition during middle childhood as well as reduced birth weight (**Tables 2-3)**. That these associations were robust to the inclusion of potentially confounding variables increases the plausibility that prenatal cannabis exposure may independently contribute to the development of behavioral problems in children. In contrast to increasingly permissive attitudes surrounding marijuana use among pregnant mothers,^41^ and recommendations by dispensaries to use marijuana to combat pregnancy-related nausea,^42^ these findings align with recent recommendations by the Surgeon General^14^ and raise concern surrounding the potential impact of in utero exposure to marijuana on mental health and cognitive outcomes in children.

All studied outcomes, except gestational age at birth, were associated with prenatal exposure before and after maternal knowledge of pregnancy (**Table 2**); however, only associations between prenatal exposure post-maternal knowledge of pregnancy and child psychopathology, cognition, and birth weight were robust to covariate inclusion (**Table 3**). There are several potential non-mutually exclusive explanations for these findings. *First*, endocannabinoid system ontogeny and signaling may play a role. Endocannabinoid type 1 receptors (CB_1_Rs), which non-human animal data suggest are critical for THC’s impact on the developing brain,^43^ are not known to be expressed before the equivalent of 5 to 6 weeks gestation in humans.^9^ As such, independent effects of cannabis on behavioral outcomes in children may only arise when sufficient CB_1_Rs are present in the fetus, which may not occur until after many women learn they are pregnant (**Table 3**).^c^ *Second*, use despite knowledge of pregnancy might represent a more severe form of cannabis use, indicative of higher and more frequent prenatal and potential post-natal exposure (e.g., during breastfeeding) consistent with a dose-response relationship that may reflect variability in liability and/or potential causal influence.^45^ *Third*, sustained use during pregnancy may reflect a genetic predisposition to the observed negative outcomes, and the resulting associations may simply reflect pleiotropy.^46,47^ However, controlling for maternal behavior and genetic susceptibility did not eliminate associations (**Supplemental eTables 2-3, 15**). *Fourth*, associations may be attributable to an unmeasured common variable (e.g., paternal germline exposure to cannabis)^48^ or an alternate derivation of an included confounder.^d^

Self-administration of cannabis, especially when heavy and early, has been strongly associated with increased psychopathology, particularly psychosis.^49^ In contrast to acute THC psychotomimetic effects,^50^ mounting evidence supports common genetic liability as a major contributor to this relationship,^39,47,51-53^ though evidence also supports potential causal effects of psychosis liability on cannabis use,^39,47^ and vice versa.^54,55^ Consistent with prior work,^21^ we find that child psychosis liability (i.e., PLEs and thought problems) is increased among children prenatally exposed to cannabis. Critically building upon this literature, this relationship was only robust to potential confounds when exposure occurred following maternal knowledge of pregnancy. That the relationship with PLEs remained after accounting for family history of psychosis as well as child polygenic risk for schizophrenia, educational attainment, and cannabis use, raises the intriguing possibility that associations between prenatal exposure and psychosis risk may not be entirely attributable to common SNP genomic liability, at least as indexed by polygenic scores derived from the largest current GWASs that approximate the child outcomes under study (but see also **Limitations** below). Indeed, putative mechanisms underlying the elevated likelihood of psychosis in children prenatally exposed to cannabis may be distinct from those associated with self-administered cannabis use. For instance, while the endocannabinoid system primarily functions as an on-demand retrograde neuromodulatory system through CB_1_R signaling throughout life, CB_1_Rs during the prenatal period are ubiquitously expressed in neural progenitors and contribute to neural migration, axonal elongation, and synaptic formation.^9^ Although such proposed mechanisms would be consistent with neurodevelopmental theories of psychosis,^56^ the associations we observed with gross brain morphology metrics were not robust to covariate inclusion. It remains possible that neurodevelopmental differences (e.g., synaptic formation) may not be detectable using MRI, may be regionally specific, or may not emerge until later developmental stages.

In addition to replicating the well validated association with reduced birth weight,^13^ children prenatally exposed to cannabis following maternal knowledge of pregnancy had elevated internalizing, externalizing, attention, and social problems, as well as reduced cognition. As has been shown for the increased likelihood of tobacco smoking during pregnancy in women with ADHD and the confounding of consequent associations with offspring ADHD,^57-59^ women with externalizing features (which include cognitive differences) might be more likely to continue using cannabis during their pregnancy. Genetically informed designs (e.g., sibling crossover design where non-twin siblings are discordant for prenatal exposure)^60,61^ provide one mechanism for identifying such familial sources of confounding.^62,63^ Notably, prenatal exposure to alcohol and tobacco were also related to offspring psychopathology and cognition, but these associations were predominantly with exposure prior to, as opposed to following, maternal knowledge of pregnancy and observed inconsistently relative to associations with cannabis exposure (**Supplemental eTables 2-3**). The lack of association with alcohol or tobacco use subsequent to knowledge of pregnancy may indicate the more pronounced public awareness of fetal risks and obstetric oversight of the use of these substances that leads to greater reductions in use post-knowledge of pregnancy, relative to cannabis.^8,64^ Alternatively, prenatal cannabis exposure may serve as a proxy for exposure to a permissive home environment that promotes externalizing behaviors and related cognitive disengagement.^65^ In addition, factors broadly correlated with prenatal substance use, such as restricted access to health care, birth-related complications and postpartum maternal behaviors, may plausibly contribute to these associations.^66^

Some limitations are noteworthy. First, parent/caregivers may have underreported marijuana use during pregnancy; however, our estimates are consistent with other national datasets, including toxicology based prevalence estimates.^67^ Second, although the ABCD study represents the largest integrative study of child health and substance use and is among the largest studies of prenatal exposure and child outcomes, there were a relatively small number of participants who were exposed to cannabis prenatally which reduces power. Third, marijuana concentration differs significantly between fetuses whose mothers use cannabis once per month compared to once per day.^15^ There are limited or no data on potency, frequency, timing and quantity of cannabis exposure during pregnancy in this dataset; further, the limited number of prenatally exposed individuals limits the utility of more refined analyses of cannabis use variability in this dataset.^68^ Fourth, while we were able to account for many known familial, pregnancy, and child related confounding variables, the role of unmeasured confounders cannot be discounted. Relatedly, while we account for underling genetic vulnerability using both familial history as well as polygenic scores, it is possible that the current GWASs from which PGS weighting was determined are underpowered and/or not generalizable across development and as such do not adequately represent genomic risk for the specific child outcomes under study, consistent with the lack of association between schizophrenia PGS and PLEs and thought problems (**Supplemental eTable 16-17**).

## Conclusions

Despite increasingly permissive social attitudes and the marked relaxation of legal restrictions on cannabis use,^1^ prenatal cannabis exposure, and the correlated risks that it indexes, may place offspring at increased risk for psychopathology and reduced cognition in middle childhood. In the context of increasing marijuana use among pregnant women,^2,6^ it is clear that more studies on the relationship between prenatal cannabis exposure and offspring developmental outcomes are needed to examine potential causal effects, moderating or protective factors, and biological mechanisms at play.^69,70^ Similar to the highly effective messaging surrounding the adverse consequences of alcohol and tobacco exposure during pregnancy, education regarding the potential harms associated with prenatal marijuana use is necessary. Currently, cannabis use during pregnancy should be discouraged by care providers and dispensaries.^11,14^

## Data Availability

Data came from the ABCD study 2.0.1 data release. More information about this study and how to access these data are available here: https://abcdstudy.org/

## ACKNLOWEDGEMENTS

Data for this study were provided by the Adolescent Brain Cognitive Development (ABCD) study which was funded by awards U01DA041022, U01DA041025, U01DA041028, U01DA041048, U01DA041089, U01DA041093, U01DA041106, U01DA041117, U01DA041120, U01DA041134, U01DA041148, U01DA041156, U01DA041174, U24DA041123, and U24DA041147 from the NIH and additional federal partners (https://abcdstudy.org/federal-partners.html). Authors received funding support from NIH: Dr. Hatoum (T32-DA007261), Dr. Johnson (F32 AA027435), Dr. Karcher (MH014677), Dr. Carter (R01-DA046224), Dr. Rogers (R01-DA046224), Dr. Agrawal (5K02DA32573; R01-DA046224), Dr. Barch (R01-MH113883; R01-MH066031; U01-MH109589; U01-A005020803; R01-MH090786), Dr. Bogdan (R01-AG045231, R01-HD083614, R01-AG052564, R21-AA027827, R01-DA046224). RB, SEP, JF, and ALM developed the research questions. SEP, JDF, AH, EJ, RB conducted analyses. SEP and RB drafted the manuscript. AH, IH, JDF, EJ, ALM, EB, YQ, NRK, EBC, CER, AA, DMB provided critical revision of the manuscript for important intellectual content. SEP, AH, JDF, EJ, and RB had full access to all data and take responsibility for the integrity of the data and accuracy of analyses. Conflict of interest disclosures: No disclosures were reported. Data on birth weight for polygenic risk scores has been contributed by the EGG Consortium using the UK Biobank Resource and has been downloaded from www.egg-consortium.org. We would like to acknowledge all participating groups of the International Cannabis Consortium, and in particular the members of the working group including S. Stringer, C. Minica, K. Verweij, H. Mbarek, E. Derks, N. Gillespie, and J. Vink.

## Footnotes

a All analyses were rerun excluding parent/caregiver respondents who were not the mother; all results and conclusions remain the same; **Supplement**).

b Cornell University was an original collection site that collected data from 34 participants, before being moved to Yale University. ABCD documentation reports 21 data collection sites and does not list Cornell; our analyses nested data based on 22 data collection sites, including the original Cornell site.

c It is possible that exposure before this time might not directly impact fetal brain development, though it remains possible for indirect effects to arise indirectly through endocannabinoid receptor expression in the placenta.^44^

d Notably, accounting for scores from a 4-item assessment of psychotic-like experiences in mothers (**Supplement**) as opposed to familial history of psychosis, which has a low endorsement rate, does not alter the significance of the association between prenatal marijuana exposure and PLEs in offspring (**Supplemental eTable 18**)

